# Improvement in Inner Retinal Function in Glaucoma with Nicotinamide (Vitamin B_3_) Supplementation: A Crossover Randomised Clinical Trial

**DOI:** 10.1101/2020.01.28.20019075

**Authors:** Flora Hui, Jessica Tang, Pete A Williams, Myra B McGuinness, Xavier Hadoux, Robert J Casson, Michael Coote, Ian A Trounce, Keith R. Martin, Peter van Wijngaarden, Jonathan G Crowston

**Author notes:** **Corresponding author:** Dr Flora Hui, Centre for Eye Research Australia, Level 7, Peter Howson Wing, 32 Gisborne Street, East Melbourne, VIC 3002, Australia. **Financial support:** Ophthalmic Research Institute of Australia (FH); Jack Brockhoff Foundation, Marian and E.H. Flack Trust – JBF Grant number 4354 – 2017 (FH); Jean Miller Foundation (JGC); Connie and Craig Kimberley Fund (IAT, JGC). Board of Research Faculty Funded Career position (Karolinska Institutet) (PAW). The Centre for Eye Research Australia receives operational infrastructure support from the Victorian Government. The funding organisations had no role in the design or conduct of this research. **Abbreviations and Acronyms:** ATP - adenosine triphosphate, CI – confidence interval, COR – coefficient of repeatability, ERG – electroretinogram, IOL – intraocular lens, IOP – intraocular pressure, IQR – interquartile range, MD – mean deviation, MLS – mean light sensitivity, NAD^+^ – nicotinamide adenine dinucleotide, NAM – nicotinamide, PL – placebo, POAG – primary open-angle glaucoma, PhNR – photopic negative response, PSD – pattern standard deviation, Q1 – lower quartile, Q3 – upper quartile, RGC – retinal ganglion cell, RNFL – retinal nerve fibre layer, VF – visual field, Vmax – saturating amplitude. **Data sharing statement:** Clinical data was collected at the Centre for Eye Research Australia in collaboration with the Royal Victorian Eye and Ear Hospital and Melbourne Eye Specialists. These data are not publicly available. Source data of graphs may be available as source files if requested. A description of the code used to analyse the electroretinogram data is described in the Methods section, and we have also published this previously (Tang et al, 2018).

## Abstract

**Importance:** Retinal ganglion cells endure significant metabolic stress in glaucoma but maintain capacity to recover function. Nicotinamide, a precursor of NAD^+^, is low in serum of glaucoma patients and its supplementation provides robust protection of retinal ganglion cells in preclinical models. However, the potential of nicotinamide in human glaucoma is unknown.

**Background:** To determine whether nicotinamide supplementation alongside conventional IOP-lowering therapy improves retinal ganglion cell function in glaucoma.

**Design:** Crossover, double-masked, randomised clinical trial. Participants recruited from two tertiary care centres.

**Participants:** Fifty-seven participants, diagnosed and treated for primary glaucoma, enrolled.

**Methods:** Participants received oral placebo or nicotinamide and reviewed six-weekly. Participants commenced 6-weeks of 1.5 grams/day then 6 weeks of 3.0 grams/day followed by crossover without washout. Visual function measured using electroretinography and perimetry.

**Main outcome measures:** Change in inner retinal function, determined by photopic negative response (PhNR) parameters: saturated PhNR amplitude (Vmax), ratio of PhNR/b-wave amplitude (Vmax ratio).

**Results:** PhNR Vmax improved beyond 95% coefficient of repeatability (COR) in 23% of participants following nicotinamide versus 9% on placebo. Overall, Vmax improved by 14.8% [95% CI: 2.8%, 26.9%], (p=0.02) on nicotinamide and 5.2% [−4.2%, 14.6%], (p=0.27) on placebo. Vmax ratio improved by 12.6% [5.0%, 20.2%], (p=0.002) following nicotinamide, 3.6% [−3.4%, 10.5%], (p=0.30) on placebo. A trend for improved visual field mean deviation was observed with 27% improving ≥1dB on nicotinamide and fewer deteriorating (4%) compared to placebo (p=0.02).

**Conclusions:** Nicotinamide supplementation can improve inner retinal function in glaucoma. Further studies underway to elucidate the effects of long-term nicotinamide supplementation.

**Trial Registration:** ANZCTR trial ID: ACTRN12617000809336 https://www.anzctr.org.au/Trial/Registration/TrialReview.aspx?id=373001

## Introduction

Glaucoma remains the leading cause of irreversible blindness worldwide.^1^ The disease has a complex aetiology and range of risk factors but is characterised by gradual dysfunction and loss of retinal ganglion cells (RGCs) and their axons which make up the optic nerve.^2^ This slow, progressive loss of visual function hinders the translation of potential neuroprotective therapeutics from bench-to-bedside due to the extended time needed to determine treatment-induced changes in glaucoma progression rates. Age,^1^ genetics,^3^ and elevated intraocular pressure (IOP)^4^ are all major risk factors for glaucoma yet clinically available treatment strategies target only IOP-lowering, and do not directly target the neurodegenerative events at the level of the retina and optic nerve. As many patients are refractory to IOP lowering or progress to blindness despite low IOP, neuroprotective strategies that directly target RGC health and decrease vulnerability to glaucoma-related cellular stressors are urgently needed. Accumulating evidence points to the potential for visual recovery in clinical glaucoma in response to IOP-lowering.^5–7^ Recovery of visual function can occur soon after restoration of IOP and, as such, objective measures of visual function have potential as surrogate biomarkers of improved RGC health.

The photopic negative (PhNR) and positive scotopic threshold responses are parameters of the electroretinogram (ERG) that are largely generated by RGCs in the inner retina.^8–10^ RGC function has been shown to recover after acute and chronic IOP insults even after prolonged periods of functional loss.^11–13^ Capacity for recovery is reduced with advancing age^14^ and can be modified by interventions such as exercise and diet restriction.^15,16^ Using ERG protocols similar to those validated in rodent studies, we have demonstrated improvements in RGC function in humans, as early as 3-months following IOP-lowering in glaucoma.^5^ Recent work has significantly improved repeatability of the PhNR measurement, enabling detection of subtle changes in inner retinal function.^17,18^

Several lines of evidence implicate oxidative stress and mitochondrial dysfunction in ageing^19^ and RGC loss in glaucoma.^20,21^ Nicotinamide adenine dinucleotide (NAD^+^) is an essential cofactor for ATP generation in mitochondria, and for NAD-consuming enzymes including sirtuins (Figure 1), with important roles in ageing, cell senescence, and stress resistance.^22^

**Figure 1.**
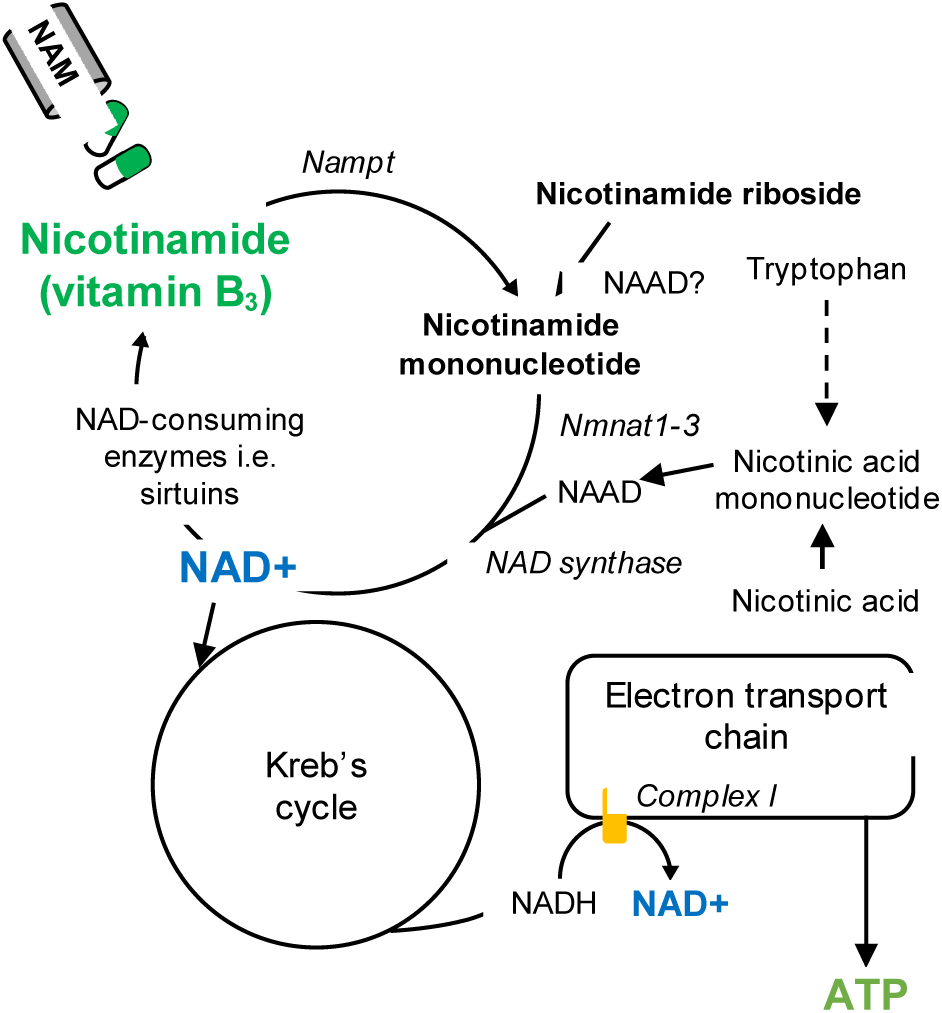
The role of nicotinamide adenine dinucleotide (NAD^+^, blue) in adenosine triphosphate (ATP) production.

The pathways through which NAD^+^ is created and salvaged, the major enzymes involved and the process in which NAD^+^ supplements, nicotinamide (green) and nicotinamide riboside, may replete NAD^+^ levels. Complex I (yellow) within the electron transport chain, where nicotinamide adenine dinucleotide + hydrogen (NADH) is oxidized to NAD and site of potential mitochondrial dysfunction in glaucoma. NAAD – nicotinic acid adenine dinucleotide.

NAD^+^ is a potent mediator of axon neuroprotection and recently, the therapeutic potential of modulating NAD^+^ metabolism has gained widespread attention in its role in ageing and neurodegenerative disease.^23–25^ A number of studies are currently investigating the effect of NAD^+^ repletion in a range of neurodegenerative disorders including Alzheimer’s disease (NCT00580931, NCT03061474) and peripheral small fibre neuropathy (NCT03912220). Recent work has indicated that patients with primary open-angle glaucoma (POAG) have reduced serum levels of nicotinamide (NAM, the amide of vitamin B_3_ and precursor for NAD^+^), indicating that systemic NAD^+^ levels may be associated with susceptibility to glaucoma.^26^ In support of this hypothesis, Williams et al^27^ demonstrated that increasing retinal NAD^+^ levels with dietary NAM or overexpression of a NAD-producing enzyme (*Nmnat1*) provided robust long-term neuroprotection of RGCs, reversed age-related transcriptomic changes, and preserved RGC function in an inherited mouse model of glaucoma (DBA/2J). These findings support the hypothesis that NAM supplementation has therapeutic potential. In addition, NAM has been safely used in a number of clinical studies with minimal adverse effects^28,29^ and, as such, NAM supplementation may be readily translated into clinical care and complement current therapies. We therefore sought to determine whether functional improvements observed with NAM supplementation in mice could be recapitulated in humans with glaucoma in an interventional study using the ERG PhNR and visual fields (VF) as main outcome measures. This study sought to determine whether NAM, a potential neuroprotectant that targets bioenergetic insufficiency in glaucoma, could lead to a detectable improvement in inner retinal function.

## Methods

### Study Design, Participants

This study was a prospective, double-masked, randomized, crossover clinical trial conducted in Melbourne, Australia between October 2017 and January 2019. The study schedule is presented in Figure 2.

**Figure 2.**
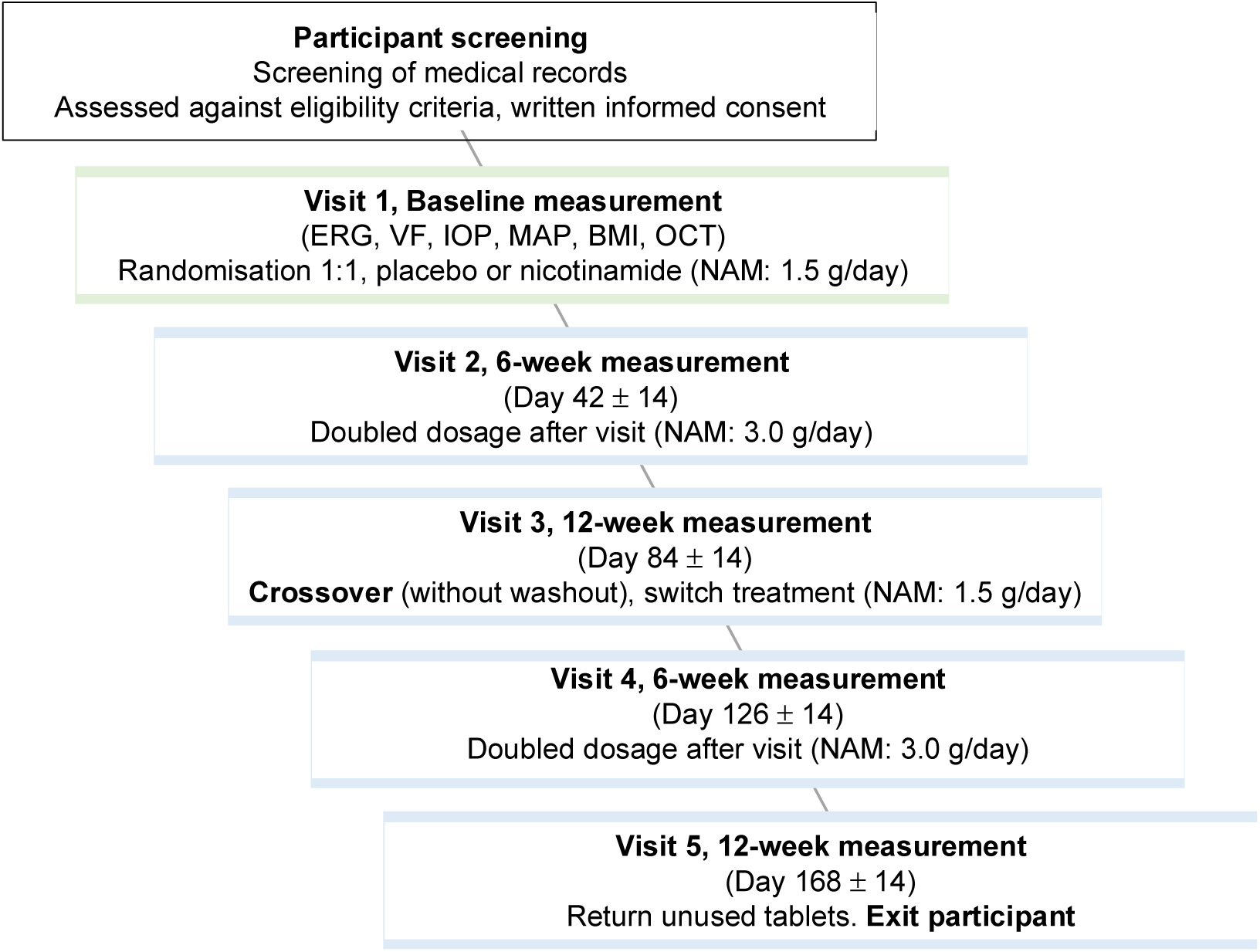
Study schedule. BMI – body mass index, ERG – electroretinogram, IOP – intraocular pressure, MAP – mean arterial pressure, NAM – nicotinamide, OCT – optical coherence tomography, VF – visual fields.

All procedures were approved by the Human Research Ethics Committee at the Royal Victorian Eye and Ear Hospital (17/1339H). Written informed consent was obtained from all participants prior to all procedures. Participants were recruited from public outpatient and private ophthalmology clinics (Royal Victorian Eye and Ear Hospital, Melbourne Eye Specialists). All testing was performed at the Centre for Eye Research Australia. Inclusion criteria included adult participants diagnosed and treated for glaucoma by a sub-specialist ophthalmologist, and with visual acuity of at least 6/18. Participants were required to have performed a reliable VF (SITA-Standard 24-2, HFA II-750i, Carl Zeiss Meditec AG, Germany) in the last 6 months, with <33% fixation losses, false positives and false negatives.^30^ Eligible participants had reproducible VF defects of at least 3 neighbouring points with a sensitivity below the age-matched normal (total deviation) with a probability of <2%.^31^ Where both eyes were eligible, the eye with the better VF was chosen.

Exclusion criteria included pregnancy/breastfeeding, allergy to NAM/niacin, history of cancer in the last 5 years (except treated basal or squamous cell carcinoma), liver disease or stomach ulcers. Ophthalmic exclusion criteria included intraocular surgery in the past 6 months (uncomplicated cataract surgery within the last 3 months) and diseases known to affect retinal function (e.g. age-related macular degeneration, diabetic retinopathy).

As this study involved high-dose NAM, an accelerated dosing protocol was used to facilitate tolerability. Participants were randomised 1:1 with randomly selected block sizes of 4, 6 and 8 using a computer-generated list to the intervention in either the first or second study period. The intervention consisted of a 6-week course of NAM (Insolar®, 0.5 g NAM tablet, Blackmores, NSW, Australia) of 1.5 g/day followed by 6 weeks of 3.0 g/day, (1.5 g twice a day, morning and evening). Placebo (PL) tablets were manufactured to match the appearance and texture of the active treatment (Pharmaceutical Packaging Professionals, Melbourne, Australia). Placebo and active treatment bottles were identical and coded with unique numerical identifiers. Participants randomised to receive placebo also doubled the number of tablets after 6 weeks to ensure they remained masked. Treatment was dispensed by the study coordinator at the conclusion of Visit 1 following capture of baseline measurements. Participants, treating physicians, certified assessors and biostatisticians were masked to treatment allocation. The NAM dosage of 3.0 g/day was based on that used in preclinical studies,^27^ other published^28^ and ongoing human trials of NAM treatment (NCT00580931, NCT03061474).

After 12 weeks, participants crossed over without washout, such that those previously on placebo treatment commenced NAM and vice versa. A washout was deemed unnecessary as any effect from NAM was assumed to be undetectable by the time of the next review visit. Treatment was taken in conjunction with any glaucoma therapies participants were already using. To monitor treatment adherence, participants were asked to bring remaining tablets to each study visit and tablets were manually counted by the study coordinator. A minimum adherence rate of 70% was acceptable, equating to no more than two missed doses a week.

### Clinical Testing

All participants were seen at baseline (Visit 1), and reviewed 6-weekly (±2 weeks). At each visit, participants underwent a standard clinical examination, including measurement of visual acuity (EDTRS letters), IOP (Icare® PRO, Icare Finland Oy, Finland), blood pressure (HEM-7322, Omron Healthcare, Japan) and slit lamp examination. Standard automated perimetry was performed on the study eye (SITA-Standard 24-2, HFA II-750i, Carl Zeiss Meditec AG, Germany) and pupils were dilated (to ≥6 mm) using 0.5% tropicamide and 2.5% phenylephrine (Bausch and Lomb, NSW, Australia). Participants were light-adapted for at least 10 minutes before photopic ERG recording. ERGs were recorded with custom-made DTL-like electrodes using silver impregnated fibre (22/1 dtex, Shieldex trading, NY, USA) and a handheld device (RETeval™, LKC Technologies, MD, USA). Reference and ground gold-cup electrodes (Grass Technologies, Astro-Med Inc., RI, USA) were placed at the temple and forehead respectively. For optimal PhNR recording, a series of red flashes (621 nm, 16 luminous energies between 0.07-12.56 cd.s/m^2^, 50 sweeps, 2 Hz flash interval,^32^ Figure 3A) on a blue background (470 nm, 10 photopic cd/m^2^) was used. Stimuli were calibrated using the ILT-1700 radiometer (International Light Technologies, MA, USA) with a photopic filter. Participants underwent optical coherence tomography (OCT) to measure retinal nerve fibre layer (RNFL) thickness (Spectralis SD-OCT, Heidelberg Engineering, Dossenheim, Germany).

**Figure 3.**
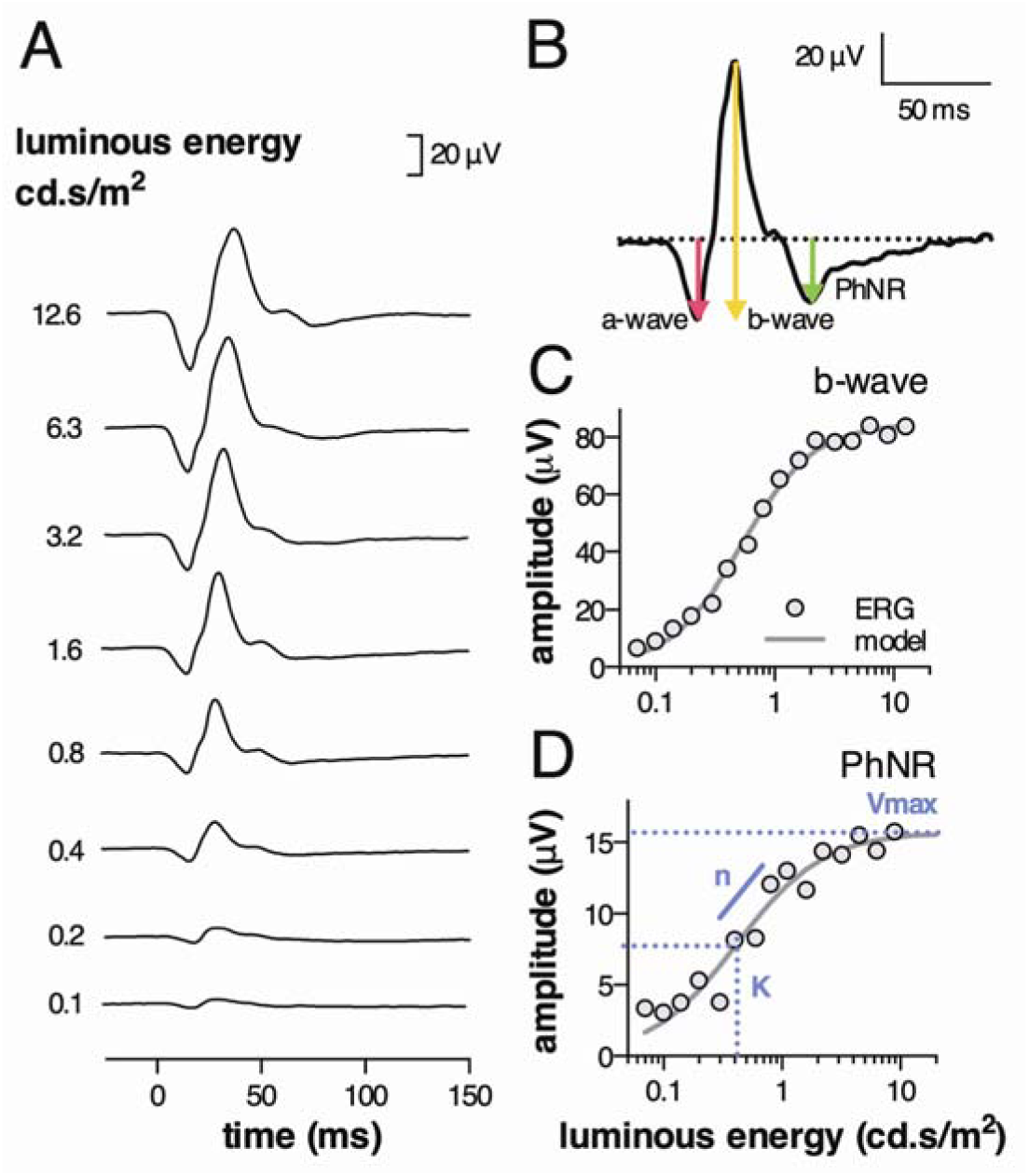
Electroretinogram (ERG) analysis method. A. Representative ERG luminance-response series showing every second luminance step, B. ERG parameters of interest, C. Representative b-wave luminance-response function data (circles) and model derived from a saturating hyperbolic function (line), D. Representative PhNR luminance-response function (circles), corresponding model (line) and model parameters: Vmax (saturating amplitude), n (slope) and K (semi-saturation constant).

**Figure 4.**
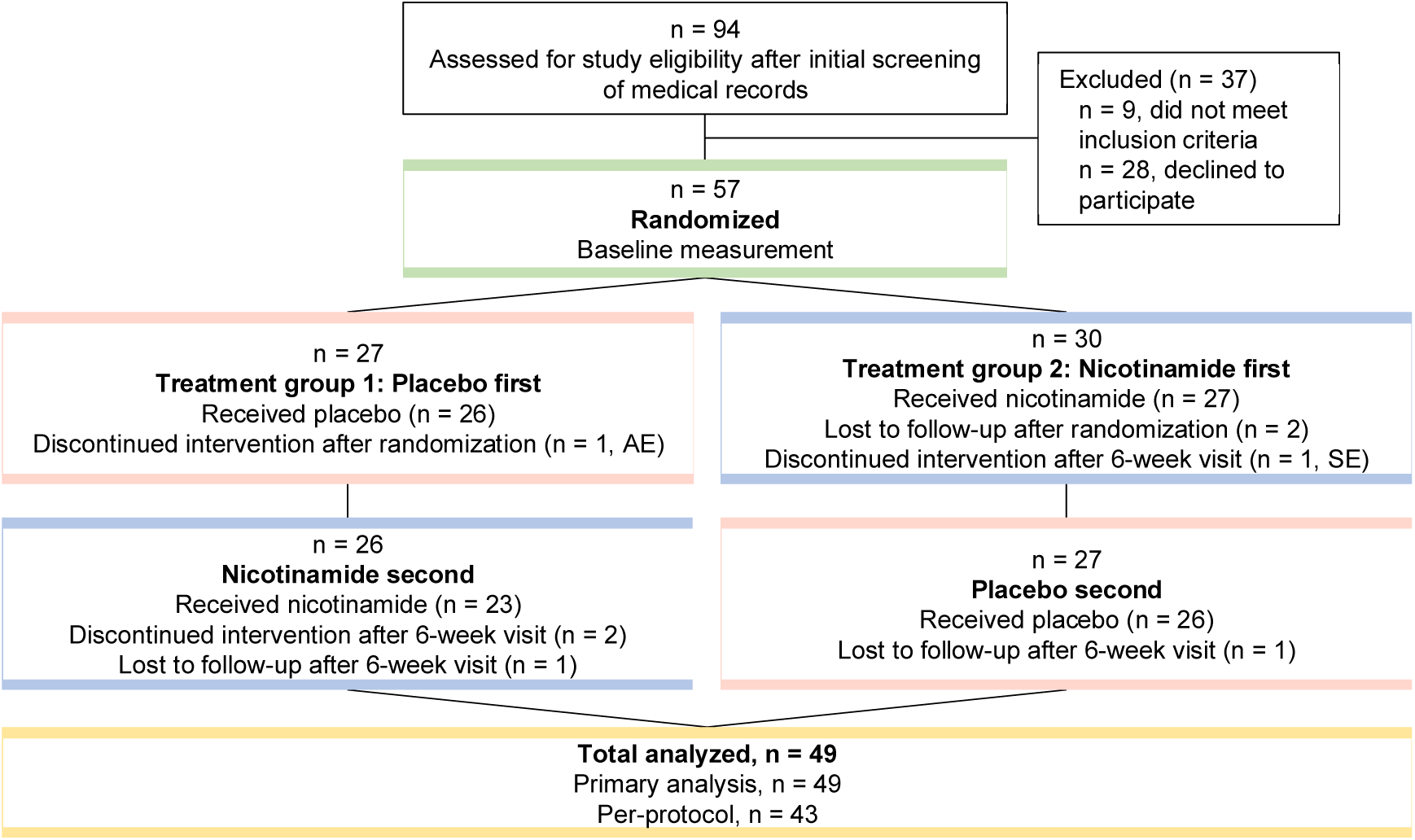
CONSORT diagram. Participants were randomised to receive placebo first or nicotinamide first. Primary analysis set defined as participants who received both interventions and attended all visits. Per-protocol set defined as a subset of the primary analysis set, excluding any participants with missing data on outcome variables (n = 6). Participants lost to follow-up (n = 4), participants discontinuing intervention (n = 4). AE – adverse event, SE – side effect.

### Study Objectives

The primary objective was to evaluate the change in retinal function as measured by ERG and VF parameters after 12 weeks of high-dose NAM supplementation compared to placebo. Specifically, changes from baseline and placebo in PhNR saturating amplitude (PhNR Vmax), PhNR/b-wave ratio (Vmax ratio) and VF indices (mean deviation, MD; pattern standard deviation, PSD; mean light sensitivity, MLS) were analysed. Secondary objectives included changes to RNFL thickness, IOP and mean arterial pressure (MAP).

### Data analysis

ERG waveforms were processed as described previously.^18^ In brief, custom-written Matlab® scripts (R2018b, Mathworks, MA, USA) were used to process raw ERG traces. ERG waveforms were processed by an assessor masked to participant characteristics, treatment group and timepoint of measurement. First, a bandpass filter (0.3 – 300 Hz) was applied. The raw data was detrended with a 3^rd^ order polynomial, which was shown to provide the most robust PhNR signal.^18^ Amplitudes and implicit times of the a-wave, b-wave (from a-wave trough to b-wave peak) and PhNR (minimum from baseline to trough) were extracted.

Amplitudes of the b-wave and PhNR (Figure 3B) across the luminance series were modelled using a saturating hyperbolic function,^33^ defined as V(I) = Vmax*I^n^/(I^n^ + K^n^), where V (μV) is the amplitude as a function of luminous exposure (I), Vmax the saturating amplitude (μV), K the semi-saturation constant (1/K is the sensitivity) and n the slope (Figure 3C-D). For the PhNR, n was fixed at 1.2 (consistent with our pilot data, and similar to others).^34,35^ Further, the PhNR Vmax was analysed as a ratio to the b-wave Vmax (Vmax ratio) to account for any changes to the b-wave between sessions.

In addition to VF MD and PSD, the mean light sensitivity (MLS) was calculated as the average of all perimetric sensitivity values on a 24-2 visual field (excluding one point above and below the blind spot). Visual field parameters are conventionally expressed in logarithmic decibel (dB) scale, however, these were converted to a linear scale (1/lambert) where, *dB* = 10 · log(l/*lambert*) before correlating with other measures.^36,37^ The proportion of participants with a ≥1 dB change in MD and PSD in NAM and placebo groups were determined and compared using Fisher’s exact test.

### Statistical Analysis

Data from one eye per participant were included in the analyses. Prior to study commencement, sample size was calculated based on results of previous work investigating short-term changes in PhNR Vmax ratio following IOP reduction (under review). Assuming an attrition rate of 20%, it was estimated that 48 eyes were required to achieve 80% power to detect a difference between treatment periods with an effect size of 0.56 for Vmax ratio and two-sided hypothesis test with an α level of 0.05.

The primary analysis set included participants who attended all study visits. Statistical analyses were conducted with the statistician masked to intervention using Stata (SE version 15.1, Texas, US). Treatment adherence was compared between randomisation groups using the Wilcoxon rank-sum test. As a washout period was not utilised, a pre-test for carryover effects was conducted for each parameter.^38^ The within-participant sum of values for each parameter was calculated by adding change from baseline values observed under placebo to those under NAM and compared between randomisation groups using two-sample t-test. To test for a period effect, the within participant difference (NAM-PL) was compared between treatment groups using linear regression, adjusting for baseline values of each parameter. The treatment effect for each parameter was assessed following the high-dose period by examining the magnitude and direction of the within-participant difference via linear regression, with adjustment for mean-centred baseline values. Values were analysed in original units of measurement. If a statistically significant treatment effect was found at 12-weeks for any endpoint, then the 6-week timepoint was also assessed. Changes in PhNR parameters were also compared to the 95% coefficient of repeatability (COR) established from our previous work.^17^

Missing ERG values were associated with fatigue or persistent muscle twitch during recordings that could not be ameliorated with post-hoc signal processing. Values were multiply imputed (25 imputed datasets) using fully conditional specification (chained equations) with a univariate linear regression imputation model to reduce bias and maintain statistical power. In addition to the imputation variables, imputation models included demographics, clinical data and study group. As a sensitivity analysis, complete-case analyses were performed using the per-protocol set (n = 43) which only included participants who had non-missing data. Results are shown as mean and [95% confidence interval], unless stated otherwise.

## Results

### Participants

Among 94 participants assessed after medical record screening, 57 were randomised into the study, of which 49 (86%, 65.5±10.0 years, 39% female) completed the study. The participant flow chart is presented in Figure 2 and baseline characteristics of the participants included in the primary analysis in Table 1.

**Table 1.** Baseline characteristics for participants in the primary analysis by treatment group (n = 49)

|  | Placebo first<br>(*n* = 23) | Nicotinamide first<br>(*n* = 26) |
| --- | --- | --- |
|  | *n* (%) | *n* (%) |
| **Sex** |  |  |
| Female | 10 (43%) | 8 (31%) |
| Male | 13 (57%) | 18 (69%) |
| **Eye tested** |  |  |
| Left | 11 (48%) | 16 (62%) |
| Right | 12 (52%) | 10 (38%) |
| **Presence of IOL** |  |  |
| No | 12 (52%) | 14 (54%) |
| Yes | 11 (48%) | 12 (46%) |
| **Glaucoma diagnosis** |  |  |
| POAG | 14 (61%) | 16 (62%) |
| CACG | 5 (22%) | 3 (12%) |
| NTG | 2 (9%) | 5 (19%) |
| PXF | 1 (4%) | 2 (8%) |
| PDG | 1 (4%) | 0 (0%) |
|  | **Mean (SD)** | **Mean (SD)** |
| Age (years) | 67.01 (2.05) | 65.30 (2.14) |
| Visual acuity (logMAR) | 0.01 (0.14) | -0.01 (0.11) |
| Intraocular pressure (mmHg) | 13.6 (2.7) | 14.5 (3.3) |
| ERG b-wave fitting parameter: *n*<sup>†</sup> | 1.32 (0.16) | 1.29 (0.17) |
| ERG b-wave Vmax (μV)<sup>†</sup> | 108.87 (27.48) | 99.00 (29.75) |
| ERG b-wave fitting parameter: 1/*K*<sup>†</sup> | 1.22 (0.30) | 1.12 (0.41) |
|  | **Median (Q1, Q3)** | **Median (Q1, Q3)** |
| Mean arterial pressure (mmHg)<sup>†</sup> | 92.17 (85.00, 96.00) | 95.67 (88.17, 98.67) |
| ERG PhNR fitting parameter: 1/*K*<sup>†</sup> | 2.40 (1.24, 3.64) | 1.64 (1.27, 2.68) |
| ERG PhNR Vmax (μV)<sup>†</sup> | 15.30 (11.10, 20.58) | 11.39 (9.31, 13.49) |
| PhNR Vmax/b-wave Vmax ratio<sup>†</sup> | 0.14 (0.11, 0.20) | 0.12 (0.10, 0.14) |
| Visual field MD (24-2) dB | -5.46 (-7.20, -1.10) | -4.51 (-8.37, -1.84) |
| Visual field PSD (24-2) dB | 6.77 (2.37, 9.85) | 5.89 (3.06, 9.28) |

<sup>†</sup>Missing values in Groups 1 and 2, respectively: ERG b-wave fitting parameter: *n*, maximum amplitude and fitting parameter: 1/*K* (*n*=2 and 3), mean arterial pressure (*n*=5 and 2), ERG PhNR (μV), fitting parameter: 1/*K* (*n*=3 and 3), PhNR Vmax/b-wave Vmax ratio (*n*=3 and 3). ERG – electroretinogram, IOL – intraocular lens, MD – mean deviation, NTG – normal tension glaucoma, POAG – primary open-angle glaucoma, PDG – pigment dispersion glaucoma, PhNR – photopic negative response, PSD – pattern standard deviation, PXFG – pseudoexfoliative glaucoma, Q1 – lower quartile, Q3 – upper quartile, SD – standard deviation, Vmax – saturated amplitude.

Majority of participants had primary open angle glaucoma (POAG, 63%), then chronic angle closure glaucoma (16%) and normal tension glaucoma (12%). Adherence rates were high for both NAM and placebo, with >94% adherence to NAM, demonstrating the high tolerability of both low and high-dose NAM supplementation (Table 2). Only two participants (3%) were non-adherent on NAM and removed from analysis after failing to attend all study visits.

**Table 2.**
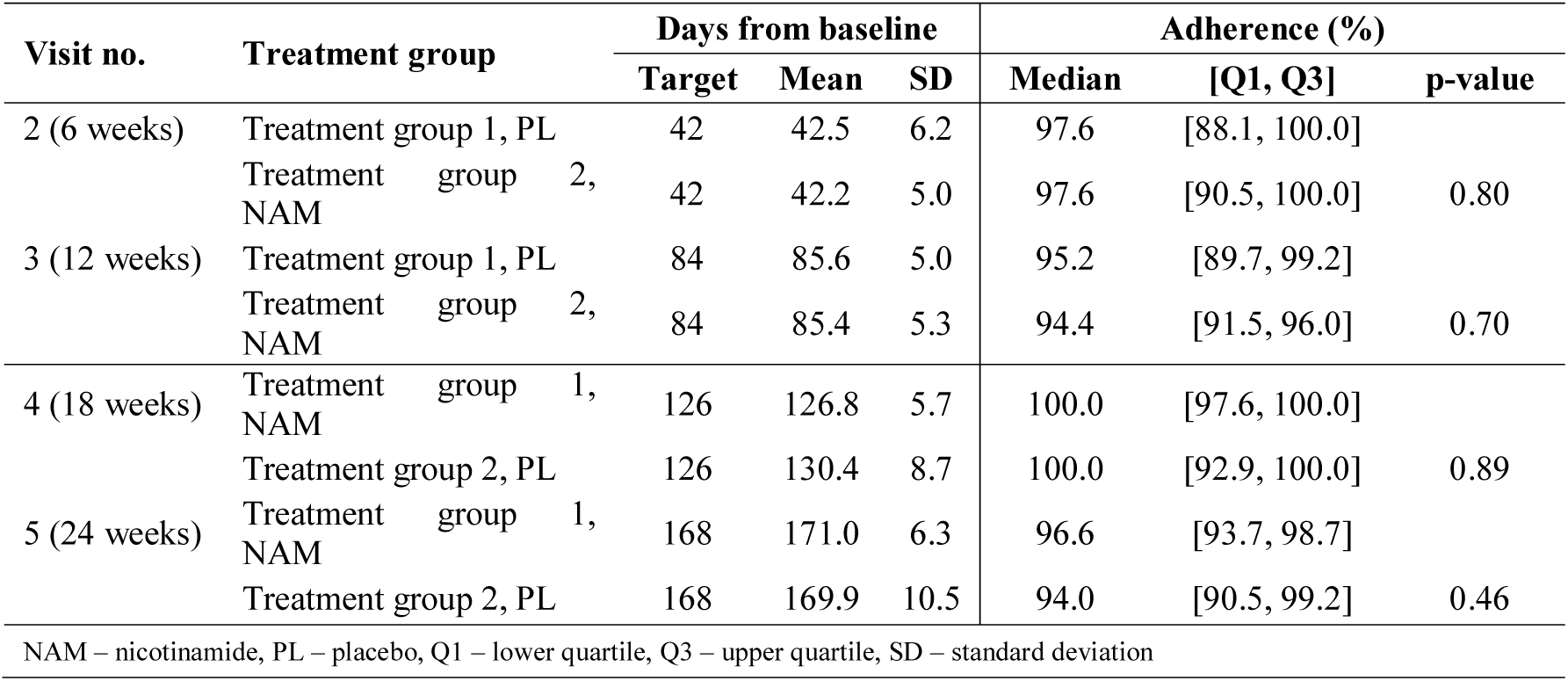
Distribution of days from baseline to follow-up for participants included in the primary analysis and adherence rates (%) at each visit. *Treatment group 1 received placebo (PL) first, Treatment group received nicotinamide (NAM) first. P-values derived from Wilcoxon rank-sum test to test for differences in adherence between groups, *n* = 49 in total*.

### Improvement in Retinal Ganglion Cell Function Following Nicotinamide Treatment

At the 12-week visit, significant improvements in inner retinal function were found in the high-dose NAM arm, with 23% (PhNR Vmax) and 21% (Vmax ratio) of the group improving beyond the respective 95% COR for each measure. Fewer participants on placebo demonstrated improvement (PhNR Vmax: 9%; Vmax ratio: 14%). Some participants deteriorated beyond the COR (9% on placebo, 7% on NAM had a reduced Vmax; 5% on placebo, 2% on NAM had a reduced Vmax ratio). A significant difference in overall treatment effect was found for both Vmax ratio (NAM-PL: 0.01 [0.002, 0.025], *p* = 0.02) and PhNR Vmax (NAM-PL: 1.35 μV [0.159, 2.551], *p* = 0.03, Figure 5A-B, 316 Supplementary Table 1). PhNR sensitivity (1/K) did not change with treatment (p = 0.41). Overall there was 12.6% [5.0, 20.2] (*p* = 0.002, Figure 5C) improvement in Vmax ratio between baseline and 12-weeks following NAM compared to 3.6% [−3.4, 10.5] (*p* = 0.30) with placebo (between group difference 9.0%, *p* = 0.03). PhNR Vmax improved by 14.8% [2.8, 26.9] *(p* = 0.02, Figure 5D) on NAM, compared to 5.2% [−4.2, 14.6] *(p* = 0.27) on placebo (between group difference 9.6%, *p* = 0.04). There was no evidence of carryover or period effects (Supplementary Tables 1 & 2) and inference was unchanged in the sensitivity analyses of the complete-case set after omitting participants with missing ERG data (Supplementary Table 2). In addition, no significant changes were noted at 6-weeks with low-dose NAM or placebo. There was no evidence of a correlation between the PhNR treatment effect and age, sex, or BMI (Supplementary Figure 1).

In addition, there was no evidence of a treatment effect on ERG measures of outer retinal function including the a-wave (photoreceptoral function, Figure 5E-F) and b-wave (bipolar cell function, Figure 5G-H) amplitudes or implicit times between NAM and placebo. No differences were found for any b-wave fitting parameters between NAM and placebo groups following the low or high-dose periods (Supplementary Table 1).

**Figure 5.**
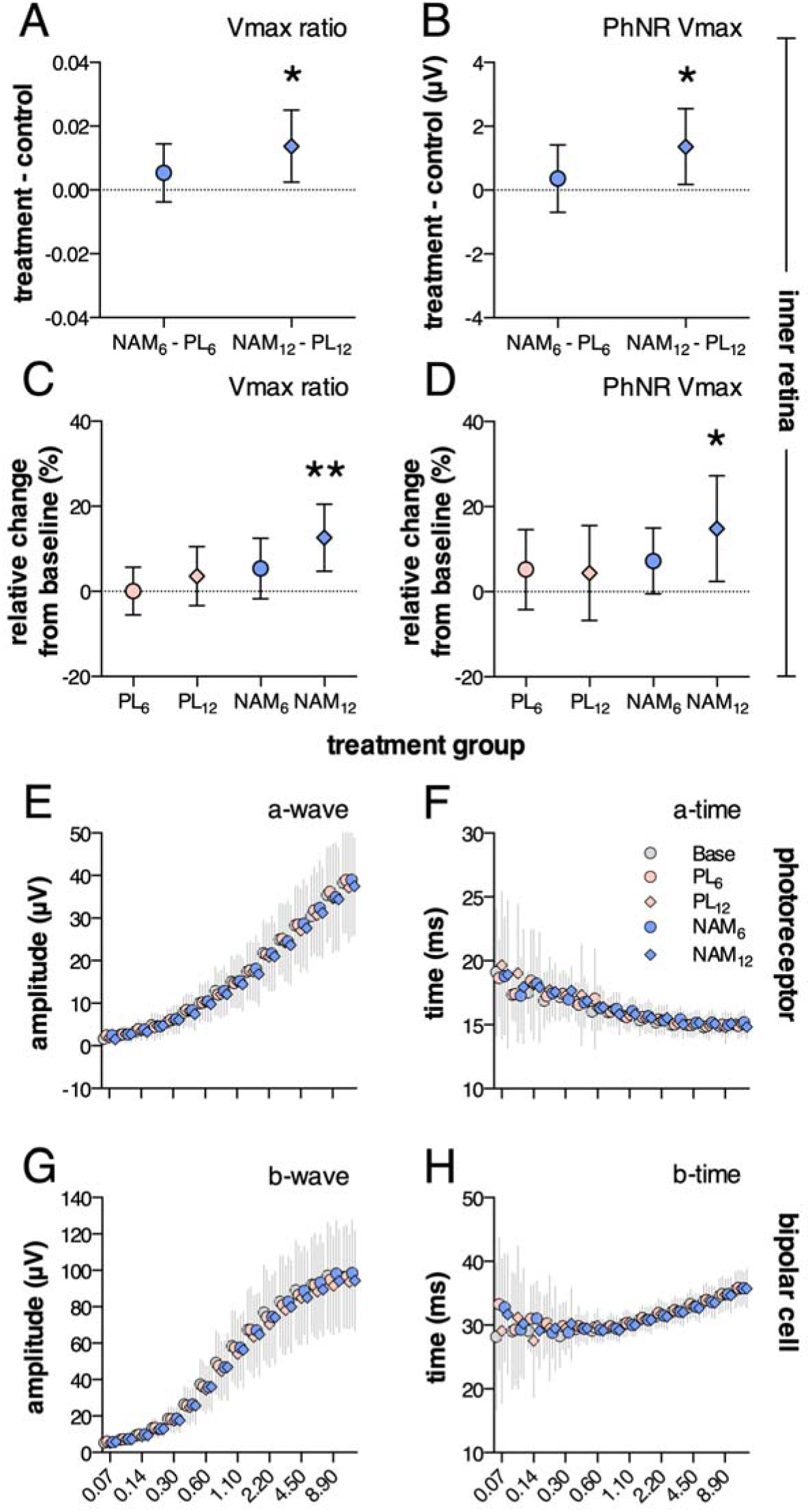
Significant improvement in the photopic negative response (inner retinal function) at 12-weeks post nicotinamide intervention. A. Difference in Vmax ratio between nicotinamide (NAM) and placebo (PL) at 6-weeks (NAM_6_ – PL_6_, circles) and 12-weeks (NAM_12_ – PL_12_, diamonds) showing significant increase at NAM_12_ (p = 0.02), B. PhNR Vmax (μV) showing a significant increase at NAM_12_ (p = 0.03), C. Vmax ratio expressed as relative change from baseline (%) for nicotinamide (blue) and placebo (pink) at 6 and 12-weeks, with a significant change at NAM_12_ (p = 0.002, one sample t-test), D. PhNR Vmax (NAM_12_, p = 0.02), E-F. Luminance-response series for the a-wave and b-wave. No significant difference in a-wave amplitude and implicit time (photoreceptoral) following placebo and nicotinamide treatment after low dose (6-weeks, circles) or high dose periods (12-weeks, diamonds). G-H. No significant change in b-wave amplitudes and implicit times (bipolar cell) following treatment; mean ± 95% CI, n = 43. Base – baseline, NAM_6_ – nicotinamide at 6-weeks, NAM_12_ – nicotinamide at 12-weeks, PL_6_ – placebo at 6-weeks, PL_12_ – placebo at 12-weeks.

### Perimetric Changes with Nicotinamide

There were no significant differences in global VF indices between treatment groups (Supplementary Table 1). After 12 weeks of NAM, average MD was no different to placebo (NAM-PL: 0.10 dB [−0.33, 0.53], *p* = 0.63, Figure 6A). Similarly, PSD reduced only slightly with NAM (NAM-PL: −0.25 dB [−0.63, 0.14], *p* = 0.20, Figure 6B). Changes in MD and PSD scores for each individual are shown in Figure 6A-B. A greater proportion of participants had an increase in MD by ≥1 dB from baseline following high-dose NAM compared to placebo (27% vs. 16%) and fewer patients taking high-dose NAM had a decrease by ≥1 dB compared to placebo (4% vs 12%, *p = 0.02*, Figure 6C). This was not evident for PSD, where a similar proportion of participants had an increase or decrease by ≥1 dB following NAM and PL (*p* = 0.61, Figure 6D). A moderate positive correlation was found between changes in PhNR Vmax and VF MD at 12 weeks (Figure 6E-F) for both placebo *(Pearson’s r* = 0.34, [0.05, 0.57], *p* = 0.02) and NAM treatment (*Pearson’s r* = 0.50, [0.24, 0.70], *p* = 0.0006). There was also evidence of a moderate positive correlation between PhNR Vmax and VF MLS (Figure 6G-H) with placebo (*Pearson’s r* = 0.37, [0.09, 0.60], *p* = 0.01) and NAM (*Pearson’s r* = 0.54, [0.28, 0.72], *p* = 0.0002).

**Figure 6.**
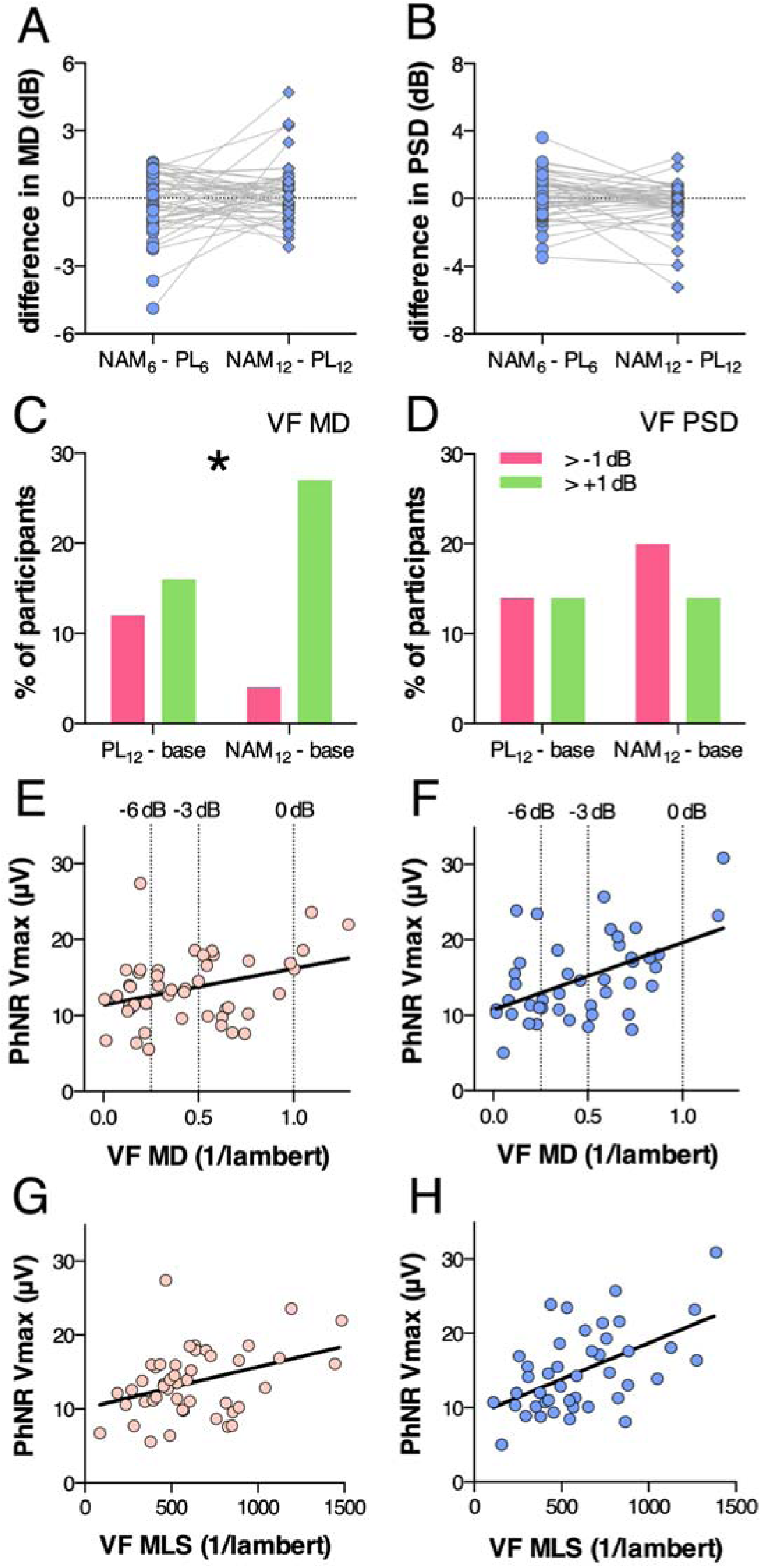
Changes in visual field (VF) parameters after treatment. A. Time-series plots of the difference in VF mean deviation (MD, dB) between nicotinamide (NAM) and placebo (PL) groups at 6 and 12-weeks for each individual, B. Difference in VF pattern standard deviation (PSD, dB) between NAM and placebo groups at 6 and 12-weeks, C. Proportion of participants (%) who demonstrated an improvement (green) or worsening (red) of VF MD by ≥ 1 dB compared to baseline for NAM and placebo groups (Fisher’s exact test, p = 0.02), D. Proportion of participants (%) who demonstrated an improvement (green) or worsening (red) of VF PSD by ≥ 1 dB compared to baseline, E. Linear positive correlation between VF MD (1/lambert) and the photopic negative response saturated amplitude (PhNR Vmax, μV) after 12-weeks of placebo (Pearson’s r = 0.34, p = 0.02). Dashed lines show where VF MD in dB units (0, −3 and −6 dB) are located, F. Correlation between VF MD (1/lambert) and PhNR Vmax after 12-weeks of NAM (Pearson’s r = 0.50, p = 0.0006), G. Correlation between VF mean light sensitivity (MLS, 1/lambert) and PhNR Vmax after 12-weeks of placebo (Pearson’s r = 0.37, p = 0.01), H. Correlation between VF MLS and PhNR Vmax after 12-weeks of NAM (Pearson’s r = 0.54, p = 0.0002); n = 49. Base – baseline, NAM_6_ – nicotinamide at 6-weeks, NAM_12_ – nicotinamide at 12-weeks, PL_6_ – placebo at 6-weeks, PL_12_ – placebo at 12-weeks.

### IOP and RNFL Thickness Unchanged

There was no evidence of a difference between placebo and NAM in IOP, MAP or VA (Supplementary Table 1). IOP was 13.8 ± 4.1 mmHg (mean ± SD) following NAM compared to 13.4 ± 2.4 mmHg following placebo (mean adjusted NAM-PL: 0.2 mmHg [−0.58, 1.003], *p* = 0.59). Overall, there was no change in RNFL thickness. At 12-weeks, the difference in RNFL thickness compared to baseline was, NAM: −0.3 ± 2.9 μm compared to placebo: 0.4 ± 2.4 μm (mean ± SD, *p* = 0.11).

### Adverse effects

In this study, high-dose NAM was well tolerated, the most common side effects being mild gastrointestinal discomfort (constipation or soft stools; 10.5%), nausea (5.3%) and headaches (3.5%). In the placebo group, 12.3% of participants reported difficulty in tablet swallowing, and 7.0% reported gastrointestinal discomfort. One participant withdrew after experiencing tinnitus whilst on placebo treatment. All symptoms resolved after participants ceased treatment.

## Discussion

We provide evidence that NAM can lead to an early improvement in inner retinal function in a significant proportion of IOP-treated glaucoma patients after 12-weeks of supplementation. The majority of individuals whose PhNR parameters improved beyond 95% COR also improved in VF MD. Overall, NAM was well tolerated with an adherence rate of >94%. As systemic NAM levels have been shown to be reduced in patients with POAG,^26^ NAM supplementation may be a convenient, safe, and cost-effective therapy in conjunction with existing IOP-lowering strategies. A longer-term study is being planned to determine whether these functional improvements are sustained and associated with delayed glaucoma progression.

There is growing evidence that impairments in visual function that occur in experimental and clinical glaucoma may recover following IOP-lowering^5,6,39–41^ or in response to the provision of bioenergetic substrates.^7,42^ A number of reports have shown short-term improvement in contrast sensitivity and VF parameters in response to IOP-lowering,^6,41,43^ and some studies indicate that these improvements in visual function are sustained for extended periods.^44^ Gandolfi et al showed significant improvements in contrast sensitivity 3-9 months after trabeculectomy, which persisted for three years.^6^ Caprioli and colleagues also demonstrated improved sensitivity on a point-wise VF analysis with long-term improvement in 44% of VF locations, five years after trabeculectomy.^44^ Studies have also provided evidence of ERG improvement following glaucoma treatment. Improvements in pattern electroretinography (PERG) were seen 3-months post-trabeculectomy^39^ and following oral acetazolamide.^40^ Niyadurupola et al demonstrated increased PhNR amplitude following a >25% IOP reduction.^5^ There is less evidence supporting visual recovery by means other than IOP-lowering. Casson and colleagues demonstrated that elevating vitreous glucose levels temporarily improved contrast sensitivity in pseudophakic individuals with POAG, in response to elevations in vitreous glucose levels.^7^ In the present study, oral supplementation with the metabolic substrate, NAM, was associated with a 14.8% [2.8%, 26.9%] improvement in PhNR Vmax amplitude and 12.6% [5.0%, 20.2%] improvement in Vmax ratio. Importantly, these effects were independent of IOP. This raises the prospect that oral NAM supplementation may serve as an adjunct to IOP-lowering therapies.

NAD^+^ depletion is observed in many tissues with advancing age^45^ and serum levels of NAM decline in aged POAG patients.^26^ The mechanisms by which NAD^+^ may improve RGC function are not known but work in animal models suggest that maintenance of mitochondrial integrity and function is a key factor.^25,27,46,47^ High-dose NAM supplementation in DBA/2J glaucoma-model mice prevented RGC soma loss, RNFL thinning, age-related transcriptional and structural changes in mitochondria and preserved RGC function as measured with PERG.^27,46^ NAM was protective as a prophylactic and therapeutic intervention in a dose-dependent manner. A similar level of neuroprotection was observed with the overexpression of NAD^+^ biosynthetic enzyme, *Nmnat1* in RGCs.^27^ A synergistic effect was found with gene therapy and NAM. Human clinical studies have not previously examined the effect of NAM supplementation on visual function in glaucoma.

NAM is the amide form of vitamin B_3_ yet is erroneously referred to as vitamin B_3_ in much of the scientific and medical literature. In these contexts, vitamin B_3_ encapsulates multiple compounds including niacin/nicotinic acid, which have known ocular and systemic side effects at high doses.^48^ However, NAM supplements are widely commercially available and are relatively safe. The most common adverse effects reported in prior clinical studies of high-dose NAM (1.5-6 g/day) were skin flushing and nausea (≤1.5%).^29^ Longitudinal studies will reveal the tolerability of long-term high-dose NAM supplementation.

We enrolled participants already treated for glaucoma with well-controlled IOP. As IOP was unchanged by oral NAM treatment, the observed effect of NAM on inner retinal function is unlikely to be mediated through changes in IOP. Instead, our findings suggest metabolic rescue of RGCs as a putative mechanism of NAM action. We hypothesise that NAM supplementation may benefit eyes with elevated IOP, which is associated with increased RGC metabolic stress and dysfunction.^2,12^ The NAM dosage used in our study was derived empirically from preclinical work and clinical trials of the supplement for other indications. Therefore, it is possible that this dose was sub-optimal and that a single dose is not ideal for all, as pharmacokinetics and pharmacodynamics will differ, for example due to age, sex, or weight.^49^ However, we did not observe a correlation between treatment effect and baseline demographics. Although a washout period was not utilized, there was no evidence of carryover effects, potentially due to the short plasma half-life of NAM in humans (3.5 hours after a dose of 25 mg/kg).^50^

For the first time, we have provided evidence that oral NAM supplementation leads to an early and measurable improvement in inner retinal function in glaucoma patients already taking IOP-lowering medication. A larger clinical trial of long-term NAM treatment is now warranted to explore whether these effects are sustained and predict a slowing in glaucoma progression.

## Data Availability

Clinical data was collected at the Centre for Eye Research Australia in collaboration with the Royal Victorian Eye and Ear Hospital and Melbourne Eye Specialists. These data are not publicly available. Source data of graphs may be available as source files if requested. A description of the code used to analyze the electroretinogram data is described in the Methods section, and we have also published this previously (Tang et al, 2018).

## Acknowledgements

Professor David Crabb for his advice on visual field analyses.

